# DAMPA: compositionally optimised probe panel design for targeted metagenomics

**DOI:** 10.64898/2026.05.15.26352859

**Authors:** Michael Payne, Kingsley King-Gee Tam, Rebecca Rockett, Kerri Basile, Rory J Bowden, Vitali Sintchenko, Jen Kok, Tanya Golubchik

**Affiliations:** Sydney Infectious Diseases Institute and School of Medical Sciences, Faculty of Medicine and Health, The University of Sydney, Camperdown, New South Wales, Australia; Centre for Infectious Diseases and Microbiology – Public Health, Westmead Hospital, Westmead, New South Wales, Australia; Centre for Infectious Diseases and Microbiology Laboratory Services, NSW Health Pathology - Institute of Clinical Pathology and Medical Research, New South Wales Health Pathology, Westmead, New South Wales, Australia; Walter and Eliza Hall Institute, Melbourne, Victoria, Australia

## Abstract

Probe-based enrichment enables highly sensitive direct-from-sample sequencing, and is being rapidly adopted for clinical metagenomics. Targeted metagenomics requires probe panels that capture hundreds of pathogens at comparable efficiency. However, as target sets expand, existing design methods often fail to account for genomic diversity, leading to overrepresentation of conserved sequences. The resulting composition bias reduces sensitivity precisely at the most clinically important and fast-evolving loci, such as antigenic regions, while increasing redundancy and sequencing cost. Here we show that panel composition bias can be minimised through biologically informed sequence compression. We implement the solution in DAMPA, a computationally efficient tool for metagenomic panel design. DAMPA uses pangenome graphs to compress conserved sequences while retaining genetic diversity, then designs probes from graph-node consensuses. In matched tests on six species, DAMPA successfully minimised composition bias, designing panels with up to 75.8% fewer probes than existing methods. In laboratory validation across four species, DAMPA-designed probes achieved sensitive and quantitative detection at Ct>30. In a highly diverse viral genome (Enterovirus A), where commercial panels retained only 7% of expected read depth, DAMPA-designed probes retained 44%, demonstrating improved robustness to sequence diversity and more uniform coverage. DAMPA enables rapid design of compositionally optimised panels that are cheaper to synthesise, while requiring less sequencing for complete genome coverage.

## Introduction

Direct-from-sample sequencing is essential for modern clinical and public health applications, including sequencing viruses and slow-growing or unculturable pathogens, wastewater surveillance and clinical metagenomics (Chiu and Miller 2019; Torres Montaguth et al. 2026; Benoit et al. 2024; You, Ni, and Shi 2024). Because shotgun (untargeted) metagenomics requires substantial sequencing effort to sensitively identify low-abundance pathogens, cost-effective targeted methods have been developed. Targeted metagenomics uses large sets of oligonucleotide probes for hybridisation capture (typically tens to hundreds of thousands per probe panel) to capture selected loci or complete genomes from hundreds of species simultaneously before sequencing (Bonsall et al. 2015; Briese et al. 2015). This approach removes the need to pre-identify or isolate the pathogen and reduces the amount of sequencing needed for pathogen detection, making it attractive for cost-effective metagenomics at scale. Targeted metagenomics is rapidly becoming a standard methodology in diagnostics, outbreak investigations, and pathogen surveillance, with over 1900 publications in PubMed, 671 just since 2025. Acceleration is in part due to the availability of very large, multi-species panels (e.g. Twist Comprehensive Viral Research Panel) (Smith et al. 2024; Boruah et al. 2023; Weeber et al. 2024; Beaudry, Bhuiyan, and Glenn 2024).

As panels grow to target a wider range of microorganisms, including viruses, bacteria, fungi and others, the number of required probes and the cost of synthesising them rapidly increase. This challenge is compounded by high genetic diversity within microbial species, which in practice greatly increases the number of probes required compared to the naïve expectation based on the length of one reference genome. Designing probe panels against a single reference per pathogen results in probes that miss regions that are highly variable across a species. Commonly used commercial panels that take this reference-representative approach, such as the Twist Respiratory Virus Research Panel and the Illumina Respiratory Virus Panel, have been correspondingly reported to have limitations in sensitivity and evenness of coverage for diverse species, such as enteroviruses (Rockett et al. 2026; Kapel et al. 2023). Viral genomes and bacterial targets with high genetic diversity are only represented by a handful of genomes, resulting in unpredictable target dropouts and limiting use of these panels for genomics in practice (Kapel et al. 2023; Burki et al. 2026).

Targeting an ever-greater number of genomes, as newer panels do, partially addresses coverage dropouts, albeit at the cost of higher probe numbers, but also introduces a critical new problem. As further reference genomes are added, conserved regions become overrepresented, resulting in highly redundant probe panels, and yet with no guarantee that the full gene content and diversity of a species will be captured. We term this problem of probe excess for conserved regions and probe insufficiency for variable regions “panel composition bias”. Panel composition bias, and the resulting loss of sensitivity for variable regions, risks missing some of the most clinically relevant antigenic and pathogenicity-related loci. These include the protein in novel coronaviruses, immune evasion genes in Cytomegalovirus and the G protein in human metapneumovirus (Li et al. 2020; Burki et al. 2026; Suarez et al. 2019). It is therefore crucial that probe panels efficiently capture the full diversity of each pathogen.

Existing probe design approaches, as implemented in tools such as CATCH and Probetools, use post hoc k-mer hashing to reduce redundancy after probe targets are already defined, but this only partially reduces composition bias: even when additional genomes are near-identical to those already in the panel (such that existing probes will suffice), redundant probes keep being added, so panel size (and bias) continues to grow with input size unrelated to underlying diversity. In an alternative approach, prior work (Bonsall et al. 2015, ve-SEQ) showed that targeting probes to a single lineage consensus instead of individual representative references works well for a single, highly diverse species (HCV). This approach is powerful because it ensures that redundant regions are covered by the smallest possible number of probes, but identifying regions to compress to consensus at the scale of a metagenomic panel has until now been infeasible.

To address the problem of generating large, metagenomics-scale panels while explicitly minimising composition bias, we developed DAMPA. In DAMPA, we draw on the developments in the field of microbial genomics, especially pangenome graphs, to identify and compress the target space for a multi-pathogen panel, at the scale of hundreds to thousands of diverse species. DAMPA collapses conserved sequences while retaining variable regions as distinct nodes within pangenome graphs, prior to probes being generated across the resulting graph structure. Describing the target space as a graph structure minimises probe redundancy and composition bias, while generating probes at metagenomic scale quickly enough for routine panel design (seconds to minutes on a laptop). We show, in laboratory validation, that DAMPA-designed panels are not only computationally efficient, but empirically more robust to diversity than a leading commercial alternative, producing less biased capture with more even sequence coverage. DAMPA is designed to run locally on a standard laptop and is available (github.com/MultipathogenGenomics/dampa).

## Materials and Methods

### DAMPA methods

DAMPA comprises three modules: *design*, *eval* and *targets*. *Design* generates a minimal probe set for a given list of target sequences. *Eval* evaluates expected performance of a probe set against a list of target sequences. *Targets* operates on the pangenome graph generated from a set of sequences to generate a minimal set of target sequences that would allow efficient mapping of any of the input sequences. These target sequences can be used for downstream analysis of targeted metagenomic sequencing data.

### DAMPA design

Input genomes are first filtered to remove undesirable sequences, including sequences with over 1% ambiguous nucleotides (i.e. N, argument maxnonstandard), sequences shorter than 100 bp (to ensure sufficient length for probe design) and sequences with a proportion of cytosine under 1% (to exclude sequences generated by bisulphite treatment). Trailing poly(A) tails longer than 5 bp are also trimmed to avoid generating probes that undesirably enrich arbitrary polyadenylated host mRNA. Database contaminants and outliers such as codon-optimised or misannotated sequences can optionally be removed (flag remove_outliers). Outliers are identified as clusters containing one sequence, or another user-defined maximum specified by the outliersizelimit argument, after clustering with cd-hit-est at 85% identity (outlierclusterident argument) (Fu et al. 2012). Input sequence sets can also be preclustered using cd-hit-est to collapse identical or highly related sequences at a sequence identity threshold (default 0.999; clusterident argument).

Filtered sequences are input to PanGraph v1.1.0 (Noll et al. 2023) with settings alpha 6.66, beta 0, identity 20, minimum length 90 by default (modifiable via DAMPA arguments pangraphalpha, pangraphbeta, pangraphident and pangraphlen, respectively). The output pangenome graph is processed to join chains of linear nodes with no branches. For target organisms or loci containing many sequences, such as global collections of viral genomes, clusters of rare or highly divergent sequences may represent misannotated genomes, passaged strains or laboratory constructs. These can be removed by specifying N, such that nodes containing fewer than N sequences will be excluded (pangraphdepth argument).

Where possible, DAMPA uses consensus sequences to minimise probe-target divergence. The rationale is that a targeted cluster of homologous sequences can be enriched by probes with a known probe-target divergence to their consensus, provided consensus-sequence identity within the cluster remains above the 80% threshold. In the case of a graph, the sequence representing each node is the majority consensus of all sequences assigned to it. These consensus sequences are then split into probes of the desired length, usually 120 bp (probelen argument), tiled end-to-end by default (probestep argument). Individual probes with more than 10 ambiguous bases (maxambig argument) or low complexity (Shannon diversity of less than 1.5, shannonthresh argument) are removed.

Finally, a modified version of probetools v0.1.11 is run using input genomes and pangenome-derived probes to identify genome regions without coverage at 80% probe-target identity and a minimum alignment length of 90 bp (arguments probetoolsidentity, probetoolsalignmin) (Kuchinski et al. 2022). Probetools then designs probes against these regions to fill any regions poorly covered by graph-derived probes.

### DAMPA eval

DAMPA eval takes a set of sequences and a set of probes. A modified version of probetools 0.1.11 is run to identify regions with missing coverage. Two plots are then generated: the first shows the number of probes at each position of each genome, and the second shows the proportion of each genome without coverage. The proportions of each genome with probe depths of 0, 1 or >1, and the mean probe depth, are reported as an average across the input sequences as well as per sequence. The total probe number and number of genomes exceeding the missing-coverage threshold (defined by argument report0covperc) are also reported.

### DAMPA targets

Downstream analysis of targeted metagenomic sequencing data using Castanet (Mayne et al. 2024) requires a database of target sequences from organisms expected to be captured based on the probes used. The genetic identity between these targets and the corresponding sample sequences should exceed 95% to ensure sensitive analysis. Corresponding target sequences can be generated from the same pangenome graph used to generate probes with DAMPA targets.

A graph component is a set of nodes that are connected in some way to each other. Each graph component falls into one of two categories: all sequences in the component belong to one species, or two or more species share at least one node.

For a species-specific component, targets can be generated by clustering the sequences within each node at 95% identity using cd-hit-est; these clusters are referred to as subclusters (Fu et al. 2012). A consensus sequence is generated for each of these subclusters. Paths through a graph are defined by the set and order of a series of nodes that recapitulates an input genome. A single target can be described as a series of subcluster consensus sequences that is defined by one or more input genome paths. All possible targets are then collected from all components in a species to generate the full target set for that species.

For a component containing more than one species, targets must be separated into individual nodes. Sequences within each node that are assigned to different species are separated and subclusters at 95% identity are generated for sequences of each species. The consensus sequence for each species-specific subcluster is then used as a target for each node. In this way species-specific target sequences from each node are generated across the component.

### Annotated genome alignment plots

Genomes of targeted organisms were first aligned using MAFFT (v7.526) with default settings. Divergence was calculated at each alignment position as the proportion of genomes not matching the consensus base. Gene annotations for HIV-1 and Enterovirus A genomes (K03455.1 and OQ091683.1, respectively) were projected onto the alignment. BLASTn (v2.16, minimum identity 85%, minimum length 80 bp) was used to determine the number of probes predicted to match each position of each genome. These matches were then used to generate the number of unique probes at each position of the alignment. Pangenome graph visualisations were generated in BandageNG (https://github.com/asl/BandageNG, (Wick et al. 2015).

### Species datasets

Genomes for Enterovirus A, JEV, Dengue and HHV4 were downloaded from NCBI if they were within 10% of the RefSeq genome size and had no more than 1% missing data (Table 1). The 4,203 Enterovirus A genomes were then clustered at 97% identity using cd-hit-est (v4.8.1) and one representative genome from each of the 768 clusters was selected (Fu et al. 2012). For *Bordetella pertussis,* a dataset of 18 genomes from a previous probe-capture study was selected (Fong et al. 2026). For HIV-1, the Group M subset of the 2021 LANL compendium was selected (Apetrei et al. 2021).

**Table 1.** Species datasets used in DAMPA development and testing.

| Species | Number of genomes | Source | subtypes | Genome size filter (Kb) | Used in |
| --- | --- | --- | --- | --- | --- |
| Japanese encephalitis virus (JEV) | 398 | NCBI virus | all | 9.9-12.1 | *In silico* analysis |
| Human immunodeficiency virus 1 group M (HIV) | 72 | Genome compendium collection from LANL* | HIV-1 Group M without recombinants | N/A | *In silico* analysis |
| Enterovirus A (EV-A) | 768 | NCBI virus + 97% identity clustering | all | 6.6-8.1 | *In silico* and validation analysis |
| Dengue virus (all 4 serotypes) | 16400 | NCBI virus | all | 9.6-11.8 | *In silico* analysis |
| Human Herpesvirus 4 / Epstein-Barr virus (HHV4, both types) | 1684 | NCBI virus | all | 155.5-190.0 | *In silico* analysis |
| *Bordetella pertussis* | 18 | (Fong et al. 2026) | Published dataset | N/A | *In silico* analysis |
| SARS-CoV-2 | 2024 | (Rambaut et al. 2020) | Pangolin lineage representatives | N/A | validation analysis |
| *Streptococcus pneumoniae* | rMLST alleles | BIGSdb (Jolley et al. 2012) | Alleles across all 53 rMLST loci linked to this species | N/A | validation analysis |
| *Enterococcus faecium* | rMLST alleles | BIGSdb (Jolley et al. 2012) | Alleles across all 53 rMLST loci linked to this species | N/A | validation analysis |

\* https://www.hiv.lanl.gov

### Tool performance comparison

DAMPA (v0.2.0) with default settings (except -t 10 for 10 processes) was used in all cases, and settings for other programs were adjusted to match DAMPA where possible. Probetools (v0.1.11) makeprobes command was used with settings -c 100, -I 85, -l 90, - L 20 -T 10, and with a batch size of 20,000 to prevent small batch size from drastically slowing probetools performance in large datasets (Kuchinski et al. 2022). CATCH (design.py) and CATCH_large (design_large.py) (v1.5.2) were both used with settings -pl 120 -ps 120 -m 18 --max-num-processes 10 --small-seq-skip 120 (Metsky et al. 2019). CATCH_large also included -e 0 so that the allowable gap between designed probes matched that of the other tools.

DAMPA (v0.2.0) eval was used to evaluate genome coverage of probe sets generated by other tools or derived from a reference genome. An additional module within DAMPA was used to select representative sequences from pangenome nodes. This module calculated pairwise distances using conserved alignment positions and selected one or more representatives so that all members of the node were captured within the design threshold (15% divergence).

Genotype III JEV genomes from the larger JEV dataset (n = 213) were used to compare the performance of DAMPA relative to Probetools and CATCH. The 213 genomes were subsampled into sets of 20 to 200 genomes in 10 replicates. To isolate the effects of increasing dataset size, subsets were nested within each replicate (i.e. the 40-genome subset contained the 20-genome subset for that replicate). Compute performance was measured on an Apple M4 Pro with 48 GB RAM.

### Validation

DAMPA was used to generate probes for Enterovirus A using the previously described dataset. SARS-CoV-2 probes were generated against a set of 2024 genomes, each representing one Pangolin lineage (Rambaut et al. 2020). For both *Enterococcus faecium* and *Streptococcus pneumoniae*, allele sequences assigned to each species were extracted from the ribosomal MLST database for all 53 loci and used to design probes (Jolley et al. 2012).

Sequencing libraries were prepared using the Twist BioSciences EF 1.0 library preparation kit, using a quarter of the manufacturer’s recommended volume. A total of 2 µl of each library (n = 96) was pooled, and the pool was captured using the Twist Target Enrichment Standard Hybridization with the custom DAMPA probes. In experiments comparing DAMPA probes and the Comprehensive Viral Research Panel (CVRP, Twist Bioscience), the same library was separately captured by each panel. Libraries were quantified using an Agilent Bioanalyzer High Sensitivity DNA Kit and a Thermo Fisher Scientific Qubit dsDNA High Sensitivity Quantitation Assay to determine the molarity of each pool. Samples were sequenced on a NextSeq 2000 using 300-bp reads. Reads were analysed using Castanet (v8) to derive target-specific deduplicated read counts (Mayne et al. 2024). Details of reads, Ct values and deduplicated read numbers are presented in Supplementary Table 1.

Amplification rate was calculated by taking the ratio of the median read depth in probe-targeted regions to the median depth of flanking genomic sequence more than 1,000 bp away.

### Publication statistics

Publication counts are from a PubMed search on 17 September 2026 with the following query: (("targeted metagenomics" OR "targeted next-generation sequencing" OR "tNGS" OR "hybrid capture metagenomics" OR "probe capture metagenomics" OR "probe-based enrichment") AND (microbial OR pathogen* OR virus OR viral OR bacteria* OR metagenomic* OR antimicrobial resistance)) NOT ("exome capture" OR "human exome" OR "cancer panel" OR "liquid biopsy" OR "oncology" OR "prenatal")

### Use of generative AI

During preparation of this manuscript, the authors used Codex (GPT-5.6 sol, OpenAI) to refine plots used in some figures. The authors reviewed and edited all AI-assisted content and take full responsibility for the final manuscript.

### Figure scripts

Scripts used in figure generation as well as additional documentation are available at https://github.com/MultipathogenGenomics/dampa-paper.

## Results

### DAMPA - consensus-based sequence collapse over a pangenome graph

DAMPA was developed to design probes that capture genomic diversity within and between genomes (Figure 1). Briefly, DAMPA represents targeted genome sequences as a pangenome graph, derives probes from the consensus sequence of each graph node and then evaluates the performance of these probes against the input genomes, generating additional probes only where gaps are identified (see Methods). The DAMPA software is readily available through Bioconda as a command-line tool. DAMPA is divided into three modules:

(1) *design* is the core probe generation process,
(2) *eval* evaluates a set of probes against a set of genomes to determine expected performance metrics, and
(3) *targets* produces a minimal set of target sequences for downstream analysis, intended for compatibility with mapping-based bioinformatics tools such as Castanet (Mayne et al. 2024). DAMPA uses the consensus of each pangenome graph node to generate probes, offering significant advantages over a representative sequence approach (Figure 1B). The probe-target identity of consensus-based probes will necessarily be higher than that of representative-based probes, reducing the overall number needed to cover the diversity of a species (Figure 1Bii). Consensus-based probes are also likely to be more robust to unsampled variation in the population as they fall closer to the common ancestor of existing clades (Figure 1Biii).

**Figure 1.**
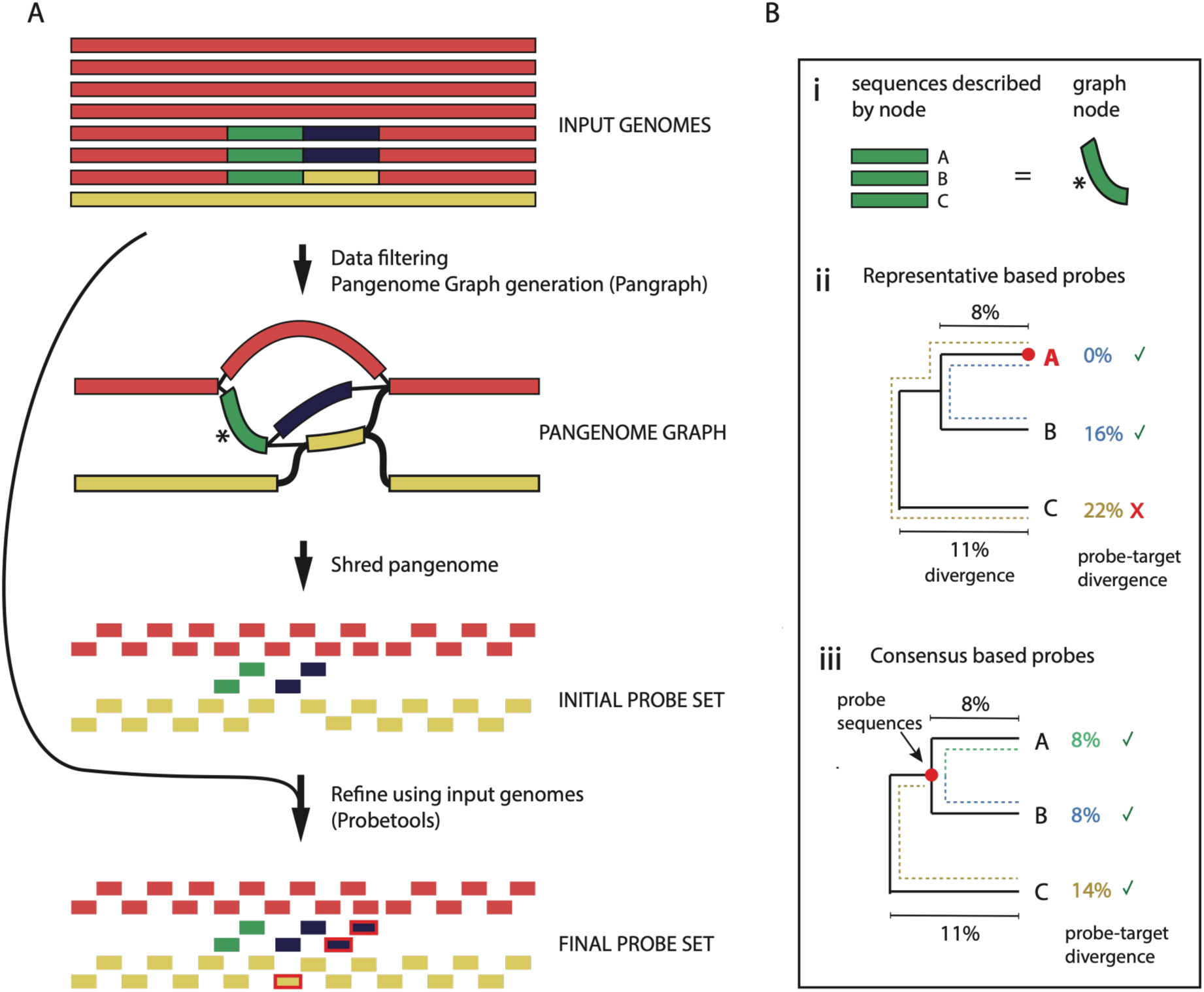
Overview of DAMPA probe design process. **A.** Input genomes are filtered for quality and input to PanGraph. The resulting graph is then processed to generate consensus sequences from each node. These sequences are then divided into 120-bp probe sequences. Input genomes are then compared against the probes to identify missing coverage and additional probes are designed. Colours represent sequences with over 80% sequence identity. **B.** Minimising probe number by using consensus sequences. The green node is composed of sequences from three input genomes (i). If probes were generated directly from the genome A sequence, then an additional set of probes from genome C would be required to capture all three sequences (ii). By using the consensus, all three sequences can be captured by a single probe set (iii).

### Naïve, reference-based panels show severe composition bias

The simplest way to design probes for a species, and to illustrate composition bias, is to divide the reference sequence into 120-bp pieces. This method is effective if all genomes and genomic regions share more than 80% identity to the reference. However, when a whole genome or genomic region falls below this identity threshold, probe-capture performance declines, resulting in missing coverage (Figure 2A) (Bonsall et al. 2015). Human immunodeficiency virus 1 (HIV-1) Group M has high within-genome diversity and Japanese encephalitis virus (JEV) has high lineage diversity, both of which require additional probes to prevent coverage loss.

**Figure 2.**
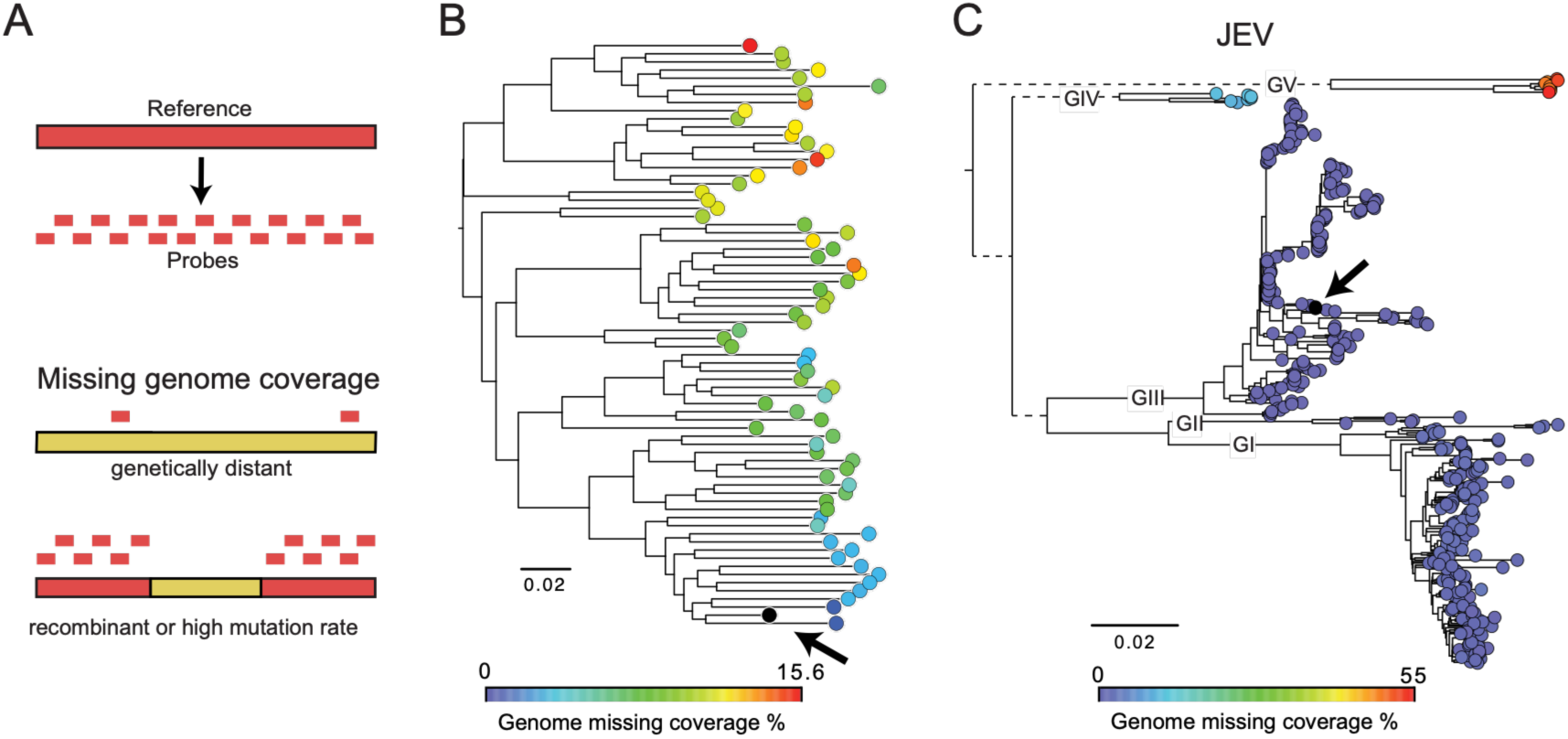
Lineage diversity causes missed probe coverage in HIV and JEV. **A.** Schematic of probes directly derived from a reference resulting in missed coverage in distant lineages or in highly diverse regions. **B.** HIV-1 Group M genomes from the LANL compendium used to construct a maximum-likelihood phylogeny. Probes were generated from the reference genome (marked by a black arrow). The proportion of each other genome in the tree not covered by one or more probes is shown by the tip colour. **C.** The same analysis as in B except for 398 JEV genomes. The five JEV genotypes are shown on their basal branches.

A single reference was used to design probes for HIV-1 Group M and JEV and the proportion of each genome that would not be successfully captured by the reference-designed probes was estimated. For HIV, the proportion with missing coverage increased across the tree as the distance to the reference increased (Figure 2B). For JEV, most genomes fell into the same large clade as the reference (which included genotypes I, II and III) and had under 0.7% missing coverage. However, genotypes IV and V had averages of 10.4% and 50.1% missing coverage, respectively (Figure 2C). In both cases the diversity of the population meant that some clades had better capture performance than others.

Genomic diversity is not uniform across a genome: differences in selection pressure and population structure lead to a wide range of evolutionary rates. For example, surface antigens that interact with the host immune system are under strong selection, leading to high diversity (*env* in HIV and VP in Enterovirus) (Figure 3A-C, Supplementary Figure 1). Reference-derived probes show composition bias because they have lower average probe-target identity across such regions. When the predicted performance of reference-derived probes was compared across the HIV genome, some regions of the env gene were predicted to capture only 2 of 72 genomes (Supplementary Figure 1A). The same bias was predicted for Enterovirus A: reference probes captured conserved regions in the 5’ UTR in 745 of 755 genomes, whereas variable regions such as VP1 were only captured in 80 of 755 genomes (Supplementary Figure 1B, marked by arrowheads). Both the lineage and within-genome diversity of any species must therefore be accounted for to avoid composition bias favouring conserved regions in a probe panel.

**Figure 3.**
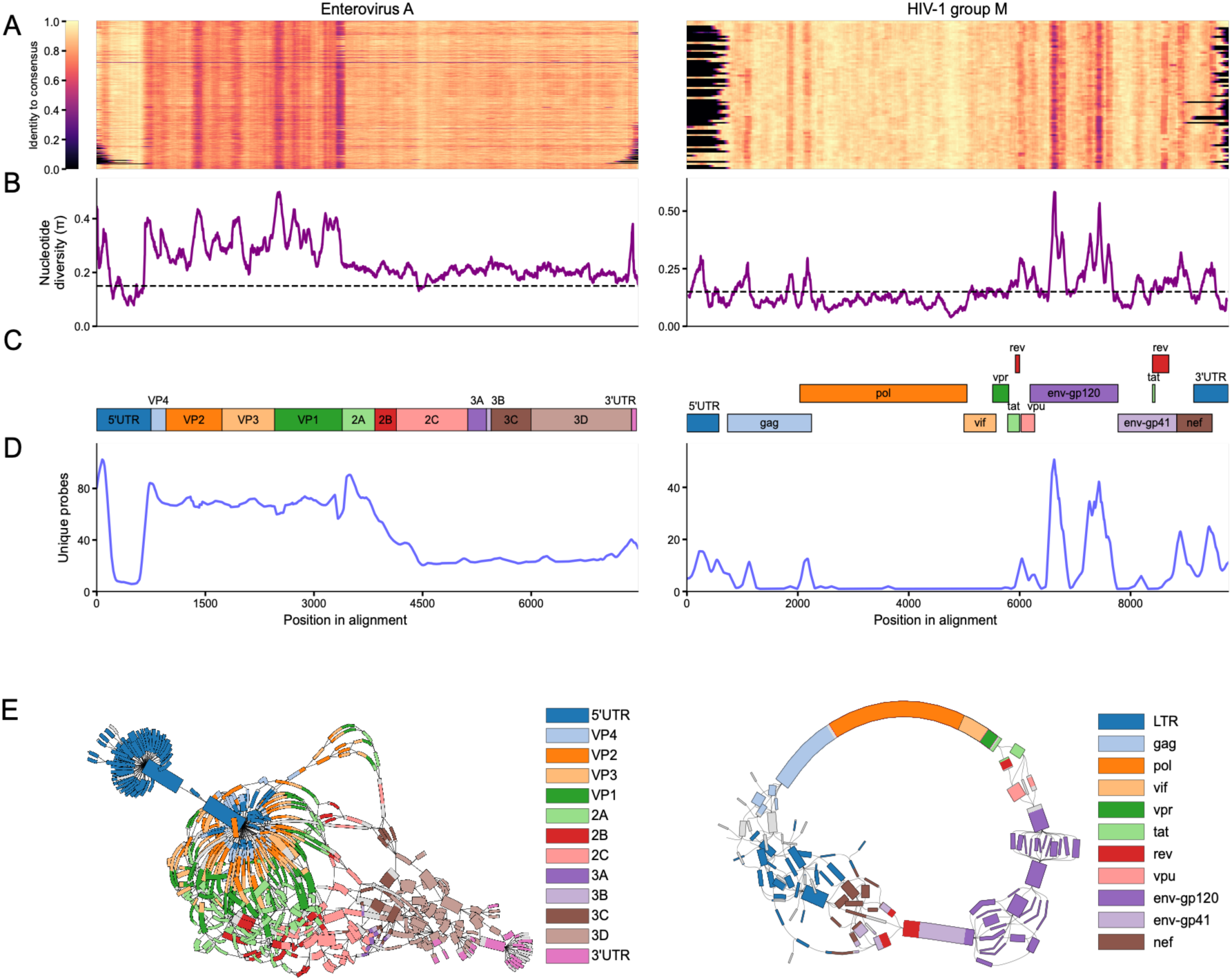
DAMPA generates probes proportional to genomic diversity via the pangenome graph. Enterovirus A genomes on the left and HIV-1 Group M genomes on the right. **A.** Heatmap of sequence conservation in each 120-bp window relative to the consensus of all input genomes in an alignment. **B.** The mean nucleotide diversity of the alignment across a 120-bp window; the dashed line indicates 15% divergence, which is the default threshold for similarity matching in DAMPA. **C.** Genome annotations from each organism projected onto the genome alignment. **D.** Number of unique DAMPA-designed probes aligning to one or more of the input genomes at each position. **E.** The pangenome graph produced by PanGraph that is used by DAMPA to generate probes. Graph colours correspond to gene annotations.

### DAMPA allocates probes in proportion to diversity, correcting composition bias

To evaluate the ability of DAMPA to remove composition bias by collapsing conserved sequences, probe sets were designed for six species (JEV, HIV, Enterovirus A, human herpesvirus 4, four dengue virus species and *B. pertussis*). The performance of these probes was evaluated relative to reference-only probe sets in comprehensive datasets for each species (Table 1 – *in silico* set). DAMPA effectively removed missing genome coverage through suppression of composition bias in all six species relative to reference-only panels (Supplementary Figure 2). The scale of the improvement in missing coverage varied from an average of 0.6% in *B. pertussis* to 64% in Enterovirus A (Supplementary Figure 2). Importantly, in all cases no more than 1% of any genome was missing coverage with DAMPA-derived probes. DAMPA successfully avoided composition bias by using the underlying pangenome graph to group conserved regions and split variable regions according to their diversity in the input genome set, generating more probes in diverse regions while retaining minimal probe numbers in conserved regions (Figures 3C and 3E). For Enterovirus A the variable VP regions generated an average of 58.9 probes per site while the conserved section of the 5’UTR region generated an average of only 6.9 probes per site (Figure 3D, left). Similarly, the most variable regions of the HIV env-gp120 gene were covered by 18 probes while more conserved regions (gag, pol, vif, vpr) were covered by a single probe (Figure 3D, right).

### Existing tools only partly correct composition bias, and don’t compact panels

DAMPA and existing k-mer based tools all collapse conserved regions when designing probes, but they use fundamentally different methods. To compare DAMPA’s graph-based approach with the k-mer hashing used by other tools, DAMPA was benchmarked against Probetools and CATCH (both standard and CATCH_large modes) using the six-species datasets used previously. Importantly, probe-target identity was required to be >85% in all tests, to provide a buffer above the well-known limit of probe binding efficiency that determines success in targeted capture in the laboratory (Bonsall et al. 2015). For all six species, all four programs generated probes at the required identity, with complete coverage of the targets (Supplementary Table 2). However, DAMPA-designed probe panels were more compact: DAMPA produced between 25.9% and 75.8% fewer probes than Probetools, between 46.2% and 63.9% fewer probes than CATCH and between 36.2% and 71.2% fewer probes than CATCH_large (Figure 4A). CATCH with default settings failed to complete for three datasets that contained larger genomes (HHV4 – 172 kbp and *B. pertussis* – 4.1 Mbp) or large numbers of genomes (Dengue – 16,400 genomes). DAMPA-designed probe sets were therefore significantly more compact than other tools, using fewer probes without compromising genome coverage.

**Figure 4.**
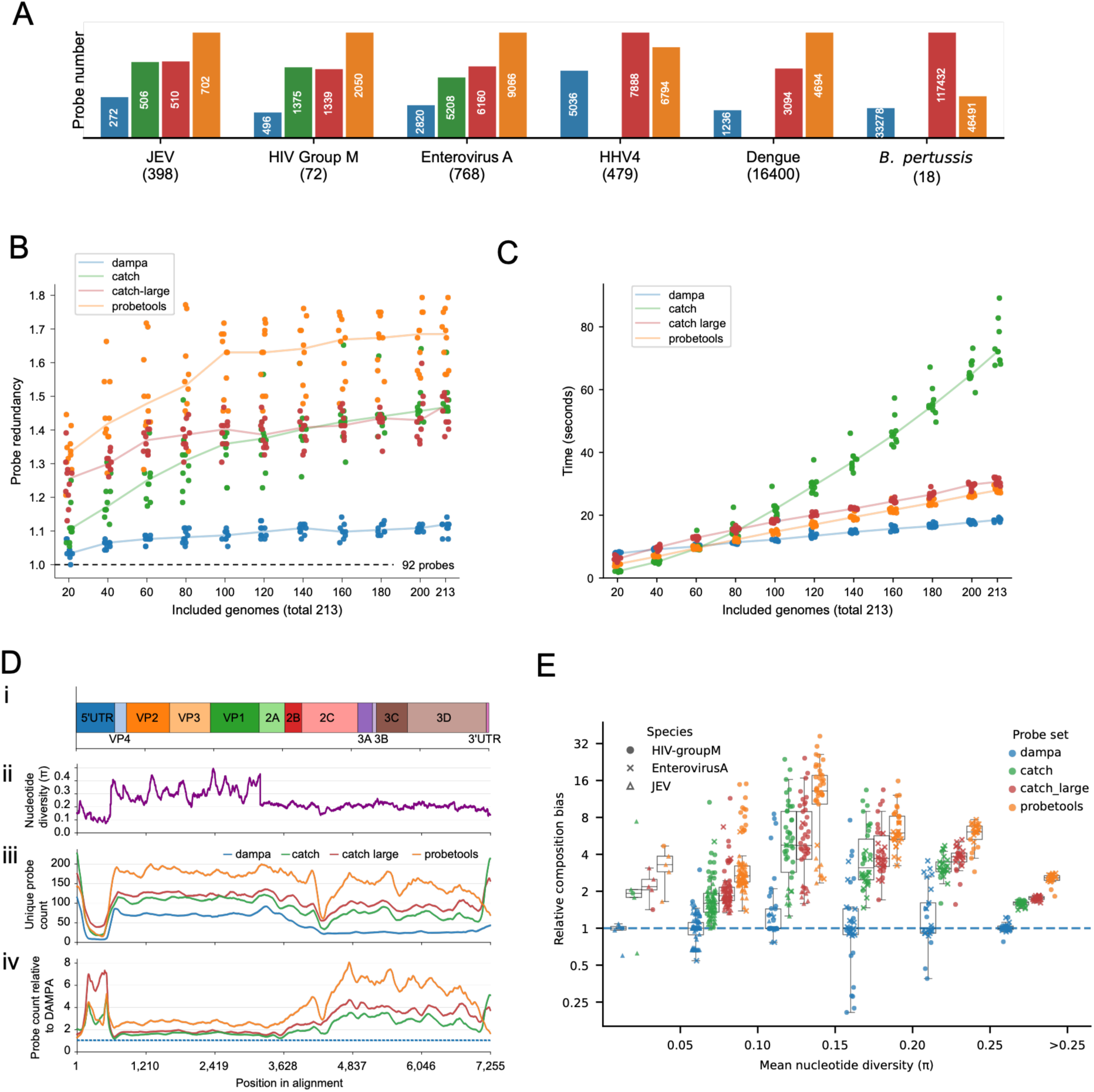
Comparison of DAMPA with CATCH and Probetools. **A.** The number of probes generated by each program in each genome set. The number of input genomes for each organism is shown beneath each plot in brackets. **B.** All genotype III JEV genomes in the input dataset (n = 213) were used to generate ten replicate subsets containing 20 to 200 genomes. Subsets were nested within each replicate. DAMPA, Probetools and CATCH were used to generate probes from each of these subsets. The theoretical optimal probe number for the genotype III genome set is 92 probes. Probe number is reported as redundancy of probes generated relative to the optimal probe number for the genotype III genome set (92 probes). **C.** The runtime for each of the subsets for each of the programs. **D.** Alignment of 768 Enterovirus A genomes. **i.** Gene annotations over the Enterovirus A alignment. **ii.** Nucelotide diversity (π) calculated per alignment position and plotted as a mean over a sliding 120bp window. **iii.** The number of unique probes predicted to bind to any genome at each position of the alignment. **iv.** The number of unique probes relative to the number of probes generated by DAMPA, used as a measure of relative composition bias. **E.** The relationship between relative composition bias and nucleotide diversity. Mean relative composition bias in each 120bp non-overlapping section of the alignment for each tool was calculated. Relative composition bias counts were binned by nucleotide diversity of the same 120bp section. All counts were normalised by the median of DAMPA counts for each bin.

K-mer-based probe-design tools tend to generate more probes with more input genomes even if there is no underlying increase in diversity. To examine this effect, all 213 JEV genotype III public genomes were used as the test dataset. These genomes have at least 90% pairwise sequence identity, meaning that their graph representation is a single node. The ideal probe set in this dataset should simply represent an end-to-end tiling of probes across the length of the genome. For JEV genotype III, the theoretical probe number needed for this tiling is 92 probes (reference length 10,976 bp / 120-bp probes). The number of probes should be unaffected by the addition of more JEV genotype III genomes, as the probe-target identity never drops low enough to require additional graph nodes. As the number of input genomes increased, DAMPA plateaued quickly, reaching a maximum of 102 probes for the full set (11% over the theoretical optimum). By contrast, CATCH and Probetools continued to add probes with the addition of more JEV genotype III genomes (Figure 4B). This tendency to add redundant probes resulted in higher final median probe counts: 135 for CATCH, 135.5 for CATCH large and 155 for Probetools, corresponding to 46%, 47% and 68% above the theoretical optimum, respectively. Additionally, DAMPA ran 3.9-fold faster than CATCH, 1.6-fold faster than CATCH_large and 1.5-fold faster than Probetools on the same 213-genome dataset (Figure 4C).

To explore the improved collapse of diversity in DAMPA, probe coverage was examined across Enterovirus A, JEV, and HIV-1 Group M genomes for all four tools (Figure 4D, Supplementary Figure 3A and B). Each 120-bp section of each alignment was treated as one data point, with mean probe depth and nucleotide diversity over that window measured. Probe counts increased with underlying population diversity as expected in all four tools (Supplementary Figure 4); however, DAMPA had fewer probes at all genomic positions except, crucially, the most diverse loci (Figure 4D). The underlying diversity at each genomic position affects the degree of improvement DAMPA shows (Figure 4D iv, Supplementary Figure 3A and B). To explore this further, relative probe number for each alignment window and tool was calculated and binned by nucleotide diversity in the same window (Figure 4E). The greatest improvement by DAMPA was near the critical probe-target identity threshold which we used for identifying regions that can be safely collapsed to consensus (0.1 < π < 0.15). Across all 120bp regions with this diversity level, DAMPA generated a median of 4.76x fewer probes than CATCH, 4.72x fewer than CATCH_large, and 13.13x fewer than Probetools. In regions with high (π>0.25) or low (π<0.1) nucleotide diversity the improvement in DAMPA averaged about 2x fewer probes than the next-most compact option.

Two complementary features of DAMPA’s algorithm contribute to its ability to generate compact panels. First, using the pangenome graph representation avoids probe redundancy in closely related genomes (Figure 4B), and second, by using the consensus rather than a representative sequence fewer probes are required to capture diverse genomes (described in Figure 1B). Below π=0.1 the improvement by DAMPA is only due to graph representation impact (Supplementary Figure 5). Above the design threshold of π=0.15, consensus improvements become dramatic, contributing half of DAMPA’s improvement over k-mer-based approaches. DAMPA thus compresses diverse regions through two synergistic effects, avoiding target redundancy with the pangenome graph and designing probes from the consensus sequences of nodes in that graph.

### Laboratory validation: DAMPA panels are sensitive and quantitative

We evaluated the performance of probes designed with DAMPA using real clinical samples. DAMPA was used to design probes to target a range of pathogens: two viruses (Enterovirus A and SARS-CoV-2), two bacterial species (*Enterococcus faecium* and *Streptococcus pneumoniae*) and four control human targets (GAPDH, COX1, STAT2 and ACTB). Because targeted metagenomics is a quantitative method that can be used to estimate pathogen load (Bonsall et al. 2020; Lin et al. 2024), we additionally evaluated DAMPA-designed probes against a serial dilution series for all four pathogens, each with independent quantification by PCR. For all four targeted organisms, DAMPA-designed probes were sensitive, with limits of detection above Ct30 (Figure 5A-D) and quantitative, with unique read counts that correlated strongly with pathogen load in dilutions in all four species.

**Figure 5.**
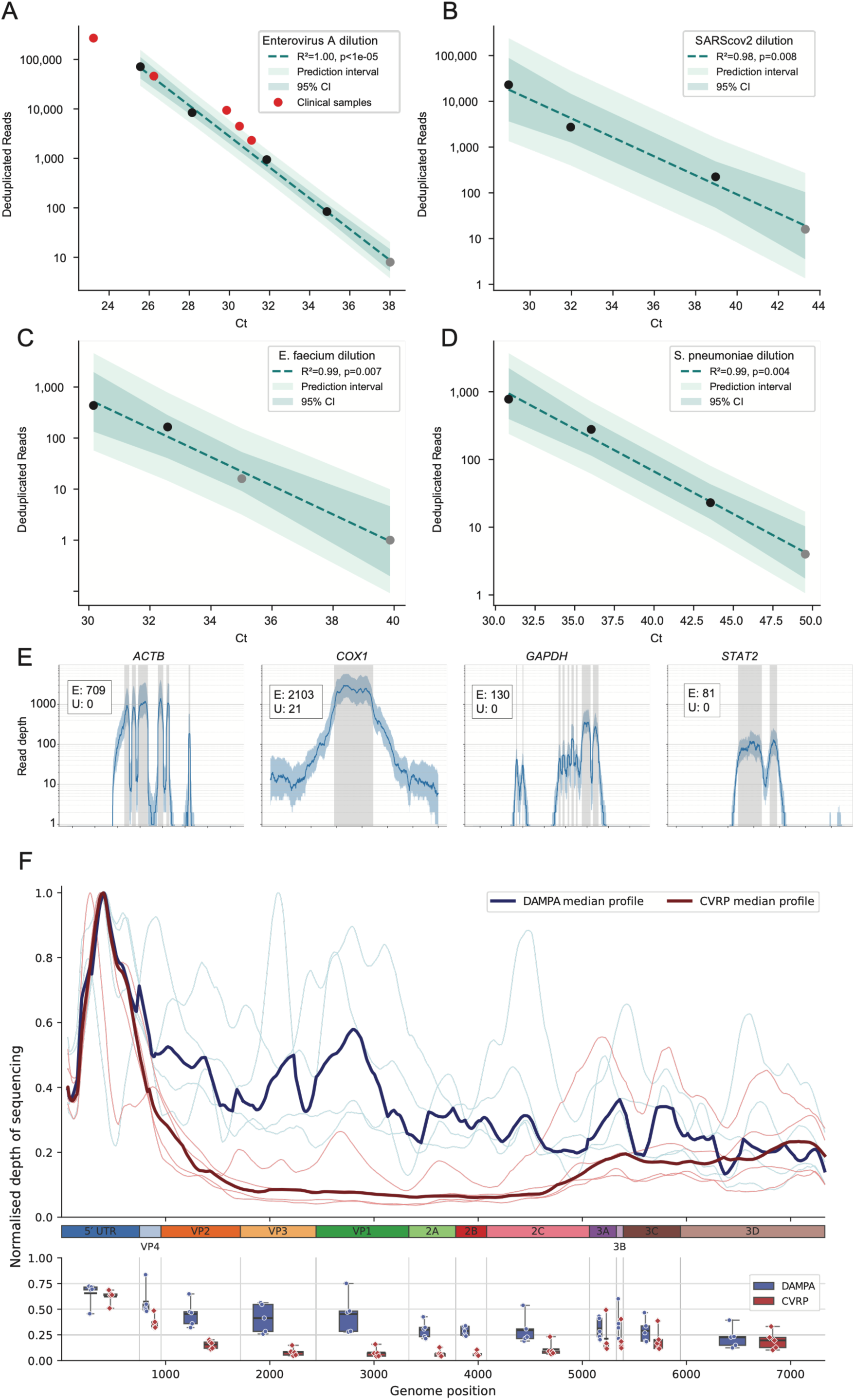
DAMPA probes are quantitative, sensitive and provide more uniform read depth in real clinical samples. **A-D.** Relationship between Ct and unique reads from targeted metagenomics using DAMPA probes. Linear regressions were performed for each plot, and R² and P value are shown. Grey points indicate imputed Ct given an assumed linear relationship of 10-fold dilutions. **A.** Enterovirus A dilutions used to generate the regression with five clinical samples of known Ct included (red points). **B.** SARS-CoV-2 dilutions. **C.** Enterococcus faecium dilutions**. D.** *S. pneumoniae* dilutions. **E.** Median read depth across the four targeted human loci; shading shows the interquartile range. *COX1* is mitochondrial, whereas ACTB, GAPDH and STAT2 are chromosomal. Grey shaded regions have corresponding probes and are expected to be enriched relative to other regions. In the insets, E and U denote the median depths of enriched and unenriched regions, respectively. Where the unenriched baseline is very low the enrichment estimates are a lower bound rather than the real enrichment number. **F.** Comparison of TWIST CVRP and DAMPA-designed probes for Enterovirus A. Read numbers for each sample at each position were collected using Castanet. Read numbers were then normalised by the maximum depth of each sample to better evaluate uniformity of coverage across the genome between methods and samples. Median normalised depth values are plotted in dark colours while each sample is plotted in lighter colours.

In addition to the dilution series, five additional clinical samples with known viral load for Enterovirus A were sequenced to assess capture in a realistic clinical sample set. These showed good agreement with the dilution standard curve (Figure 5A, red points). The observed capture performance showed that DAMPA-designed probes were highly efficient and achieved high target coverage with minimal probe redundancy. For Enterovirus A, DAMPA generated significantly fewer probes than Probetools or CATCH but demonstrated efficient capture performance even in samples with low pathogen loads.

Enrichment levels were estimated based on the human control targets to evaluate capture performance. Across all clinical samples, median enrichment was at least 709, 100, 130 and 81-fold for ACTB, COX1, GAPDH and STAT2, respectively (Figure 5E). The advantage of targeting a mitochondrial gene, *COX1,* was that sufficient sequencing depth could be generated at the remaining untargeted mitochondrial regions to estimate true enrichment above metagenomic background level. Based on this estimate, DAMPA-designed probes facilitated enrichment to 100 times above background level (median enriched depth 2103x, compared with 21x background).

### DAMPA panels are more robust to diversity: reduced composition bias translates into more uniform coverage than a major commercial panel

Probe panels should optimally capture comparable read numbers for all targeted genomic regions, regardless of differing levels of sequence diversity. Comparable enrichment ensures uniform coverage, preventing wasted sequencing effort that results from over-sequencing of conserved regions at the expense of variable ones. The performance of DAMPA probes was compared directly with that of the popular Comprehensive Viral Research Panel (CVRP, Twist Bioscience) in five Enterovirus A clinical samples. Although both CVRP and DAMPA probes over-enrich the highly conserved 5’UTR region, which is shared among the entire Enterovirus genus, capture with DAMPA probes resulted in significantly more consistent and uniform genomic coverage than CVRP (Figure 5F). Relative to the 5’UTR peak, DAMPA probes retained 43.5% read depth at the VP1 gene while CVRP probes retained only 7%. This sharp decrease in probe binding relative to the conserved 5’UTR by CVRP therefore requires greater sequencing depth for CVRP-captured libraries to generate the same level of genome-wide coverage as is obtained with DAMPA using significantly fewer reads.

## Discussion

In targeted metagenomics, the optimal oligonucleotide probe panel includes the fewest probes necessary to capture all members of a species. Given that the minimum probe-target identity for consistent, efficient capture is 80%, the genetic diversity of a species must be critically taken into account during probe design (Bonsall et al. 2015). Genetic diversity can be distributed across the genome or concentrated on certain loci due, for example, to differences in selective pressures, population structure, or recombination level. If either diversity type exceeds 20% divergence from a given genome, probes designed from that genome will lose capture efficiency, as seen in JEV genotypes IV and V. This is problematic in a surveillance or research setting, where biased results can occur if different clades are captured with differing sensitivity, favouring strains close to the genomes used to generate probes. The loss of efficient capture in highly variable regions within a genome is also problematic, as these regions are often of significant interest due to their interactions with the host immune system (e.g. *env* in HIV and VP in Enterovirus). Therefore, attempts to use targeted metagenomic data to study variation in these regions (for example, to identify vaccine escape variants) may be hampered if pathogen diversity is not accounted for.

Removing panel composition bias through the collapse of regions conserved across a species is essential for the design of efficient, effective probe panels. Several probe design methods have been developed that generate probes from all input genomes, cluster them to reduce redundancy, and then select a probe set that covers all targeted regions (Kuchinski et al. 2022; Metsky et al. 2019). However, this approach immediately splits input genomes into k-mers or probes, thereby losing the opportunity to make informed design decisions based on evolutionary similarity. As a result, each output probe is derived directly from a representative input genome, with limited opportunity for further refinement.

An alternative approach first proposed over a decade ago (Bonsall et al. 2015) showed that targeting probes to a subpopulation consensus, rather than individual representatives, can efficiently capture a diverse pathogen. Here, we show that this result generalises beyond the single-genome level, and that the consensus approach can be used to design better probe panels for metagenomics. To automate this approach for any species with any level of diversity, we leveraged recent methodological developments in microbial pangenome graphs (Noll et al. 2023). This facilitates the identification of sets of conserved regions of the genome (pangenome nodes) that can be most efficiently collapsed into a consensus at a probe design threshold.

The combination of pangenome graphs and consensus targeting in DAMPA allows the generation of more compact probe panels by two complementary mechanisms: first, by avoiding probe redundancy in conserved regions by retaining larger genomic context, and second, by allowing the use of evolutionary relationships between sequences to reduce probe number using the consensus. As targeted metagenomics panels grow in breadth, targeting ever larger numbers of highly diverse pathogens, DAMPA’s improved representation of underlying diversity can reduce costs by decreasing the number of probes required per pathogen. Consensus-based probes derive their robustness by more closely matching different lineages, implying robustness to emerging unsampled lineages. Probes designed directly from representative genomes are, by definition, further diverged from the common ancestor and are therefore likely to differ more from newly emerging lineages, potentially reducing capture efficiency. By contrast, DAMPA’s consensus-based probes more closely represent the common ancestor of a sequence set and may therefore capture a novel clade efficiently if it shares that ancestor.

We demonstrate that DAMPA-designed probes can sensitively and quantitatively enrich a variety of targets including bacterial, viral, and human targets in the same library, with comparable enrichment levels regardless of the ultimate source of the targeted sequence. Compared with the leading commercial panel CVRP, DAMPA probes generate sequencing results with more even coverage. Reduction in panel composition bias means that less sequencing is wasted on conserved regions at the expense of diverse ones, with reduced risk of coverage loss and dropouts at variable regions. DAMPA’s ability to yield more uniform coverage implies less sequencing is required to achieve the same genome coverage, reducing costs. By reducing panel composition bias, DAMPA-designed probes may therefore reduce both panel-synthesis and sequencing costs.

## Conclusion

DAMPA leverages pangenome graphs to design probe sets for targeted metagenomics that reflect the underlying diversity of the population, ensuring that rapidly evolving regions are captured with the same sensitivity as conserved regions. DAMPA probe sets were more compact and showed less composition bias than those generated by existing methods, resulting in more affordable targeted metagenomics experiments. Importantly, DAMPA-designed probes have been validated in real-world clinical samples and showed high sensitivity and improved sequencing evenness due to reduced composition bias, consequently reducing required sequencing depths and cost.

## Data Availability

DAMPA is available as an open source package that can be installed with conda, and is free for academic use. https://github.com/MultipathogenGenomics/dampa
Sequences generated as part of the laboratory validation are available from the ENA project PRJNA1466720.

## Software availability

DAMPA is freely available for academic use and can be installed with Conda. https://github.com/MultipathogenGenomics/dampa. Scripts used to generate figures are available at https://github.com/MultipathogenGenomics/dampa-paper.

## Data availability

Sequences generated as part of the laboratory validation are available from the ENA project PRJNA1466720.

## Ethics

Clinical samples and metadata were collected by the PRL at the NSW Health Pathology-Institute of Clinical Pathology and Medical Research under the Western Sydney Local Health District Human Research Ethics and Governance Committee (Project identifier: 2020/ETH02426). All data were de-identified.

---

**Supplementary Figure 1.** The relationship between nucleotide divergence and reference-derived probe performance in HIV and Enterovirus A. A. i. Genome annotations from HXB2 projected onto an alignment of HIV-1 Group M genomes (n = 72). ii. The percentage of genomes at each position that are different from the consensus. Plotted as a rolling average over a 120-bp window. iii. The number of genomes predicted to be covered by a probe derived from the HXB2 reference at each position. Plotted as a rolling average over a 120-bp window. Arrowheads mark minimum genome count positions. B. As for A except for Enterovirus A genomes (n = 755) using the OQ091683.1 genome as a reference.

**Supplementary Figure 2. Reference- and DAMPA-derived probe-set performance. A.** The proportion of each genome that is not predicted to be captured by either reference-derived (grey) or DAMPA-derived (blue) probes. Distributions are also summarised by boxplots. **B.** The proportion of genomes in each dataset that have either more than 50% missing coverage, 1–50% missing coverage or under 1% missing coverage for reference and DAMPA-derived probes.

**Supplementary Figure 3. Relationship between probe count from 4 tools and nucleotide diversity using JEV and HIV genome alignments. A.** HIV1 GroupM genomes (72) alignment **B.** JEV genomes (398) alignment. For each organism, the four panels show the following analyses: **i.** Gene annotations over the alignment. **ii.** Nucelotide diversity (π) calculated per alignment position and plotted as a mean over a sliding 120bp window. **iii.** The number of unique probes predicted to bind to any genome at each position of the alignment. **iv.** The number of unique probes relative to the number of probes generated by DAMPA, used as a measure of relative composition bias.

**Supplementary Figure 4. Relationship between probe count from 4 tools and nucleotide diversity plotted as 120bp datapoints.** Unique probe counts from each of DAMPA, CATCH, CATCH large and Probetools as well as nucleotide diversity (π) were calculated from non-overlapping 120bp alignment sections from Enterovirus A, HIV1 group M and JEV alignments. Each section was plotted for each tool against π. Tools are indicated by colour and species by data point shape.

**Supplementary Figure 5. Performance of DAMPA using representative-based rather than consensus-based probes. A.** Probes were designed against the 6 species datasets with a modified version of DAMPA that uses representative sequences from each pangenome graph node rather than the consensus (as described in figure 1B). **B.** For Enterovirus A, probe counts across the alignment are shown for the four tools and the DAMPA representative-sequence variant. **i.** Gene annotations over the alignment. **ii.** Nucelotide diversity (π) calculated per alignment position and plotted as a mean over a sliding 120bp window. **iii.** The number of unique probes predicted to bind to any genome at each position of the alignment. **iv.** The number of unique probes relative to the number of probes generated by DAMPA, used as a measure of relative composition bias. **C.** The relationship between relative composition bias and nucleotide diversity. Mean relative composition bias in each 120bp non-overlapping section of the alignment for each tool was calculated. Relative composition bias counts were binned by nucleotide diversity of the same 120bp section. All counts were normalised by the median of DAMPA consensus probe counts for each bin. **D.** A comparison of the relative contribution to DAMPA’s reduced probe count relative to the closest k-mer based tool. Difference between CATCH and DAMPA-representative relative composition bias represents the improvement attributable to avoiding probe redundancy (brown). The difference between DAMPA representative and consensus relative composition bias represents the improvement attributable to using the consensus sequence to derive probes. The contribution of these two factors is shown across different nucleotide diversity bins.

