## Supplementary Figure 1 for "DAMPA: compositionally optimised probe panel design for targeted metagenomics"

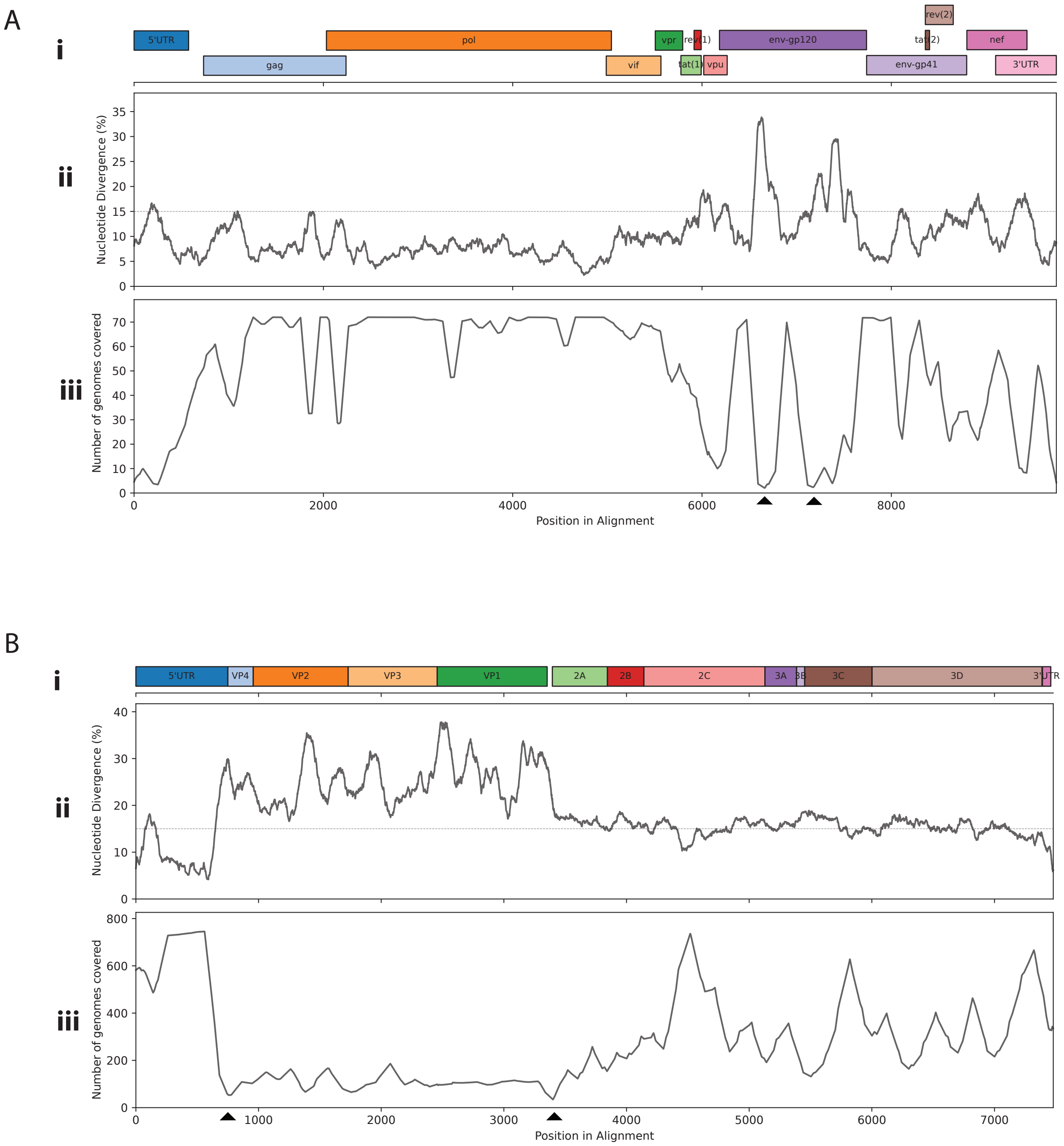


**Supplementary Figure 1.** The relationship between nucleotide divergence and reference-derived probe performance in HIV and Enterovirus A. A. i. Genome annotations from HXB2 projected onto an alignment of HIV-1 Group M genomes (n = 72). ii The percentage of genomes at each position that are different from the consensus. Plotted as a rolling average over a 120-bp window. iii The number of genomes predicted to be covered by a probe derived from the HXB2 reference at each position. Plotted as a rolling average over a 120-bp window. Arrowheads mark minimum genome count positions. B. As for A except for Enterovirus A genomes (n = 755) using the OQ091683.1 genome as a reference.
