## Supplementary Figure 2 for "DAMPA: compositionally optimised probe panel design for targeted metagenomics"

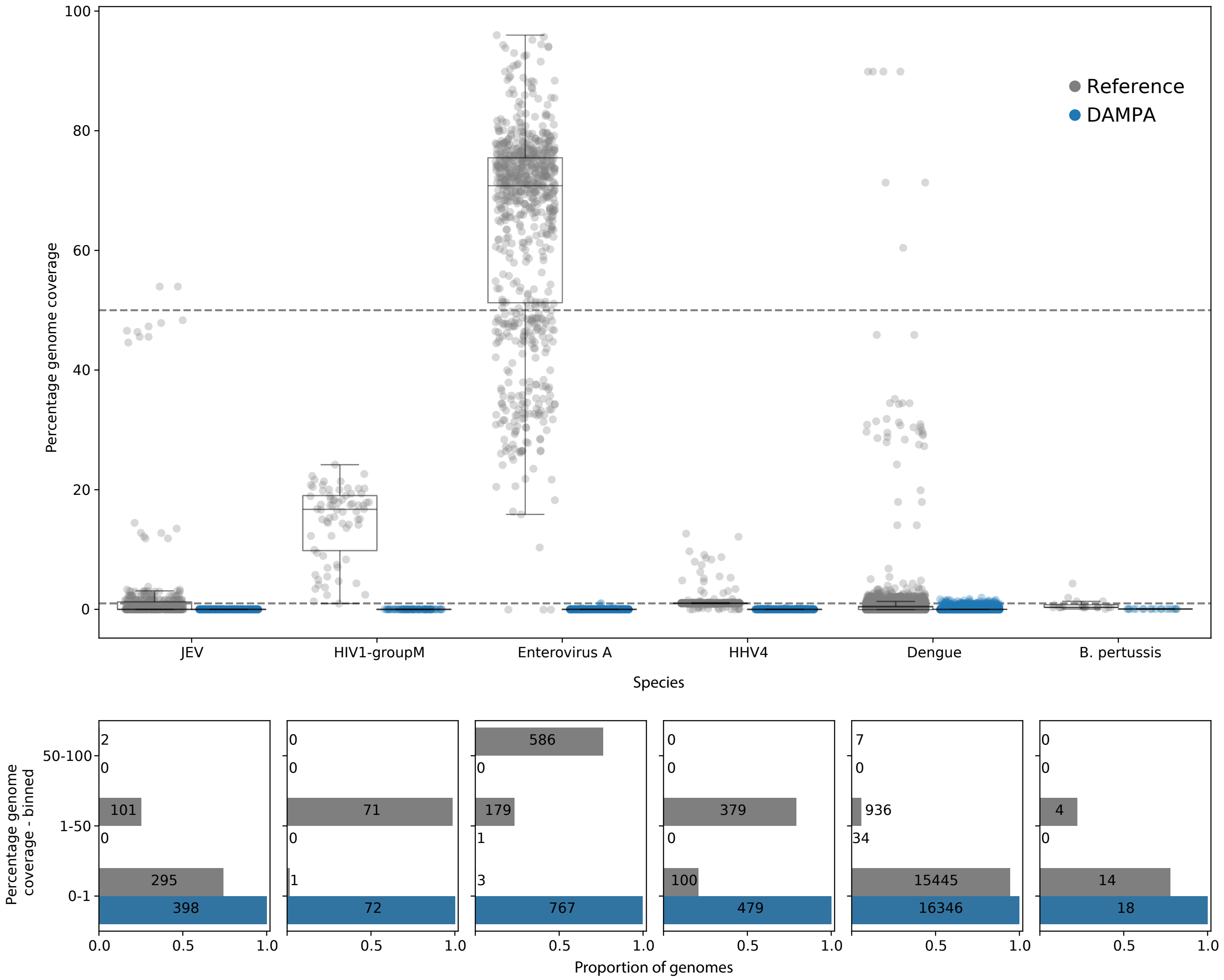


**Supplementary Figure 2.** Reference- and DAMPA-derived probe-set performance. A. The proportion of each genome that is not predicted to be captured by either reference-derived (grey) or DAMPA-derived (blue) probes. Distributions are also summarised by boxplots. B. The proportion of genomes in each dataset that have either more than 50% missing coverage, 1–50% missing coverage or under 1% missing coverage for reference and DAMPA-derived probes.
