## Supplementary Figure 3 for "DAMPA: compositionally optimised probe panel design for targeted metagenomics"

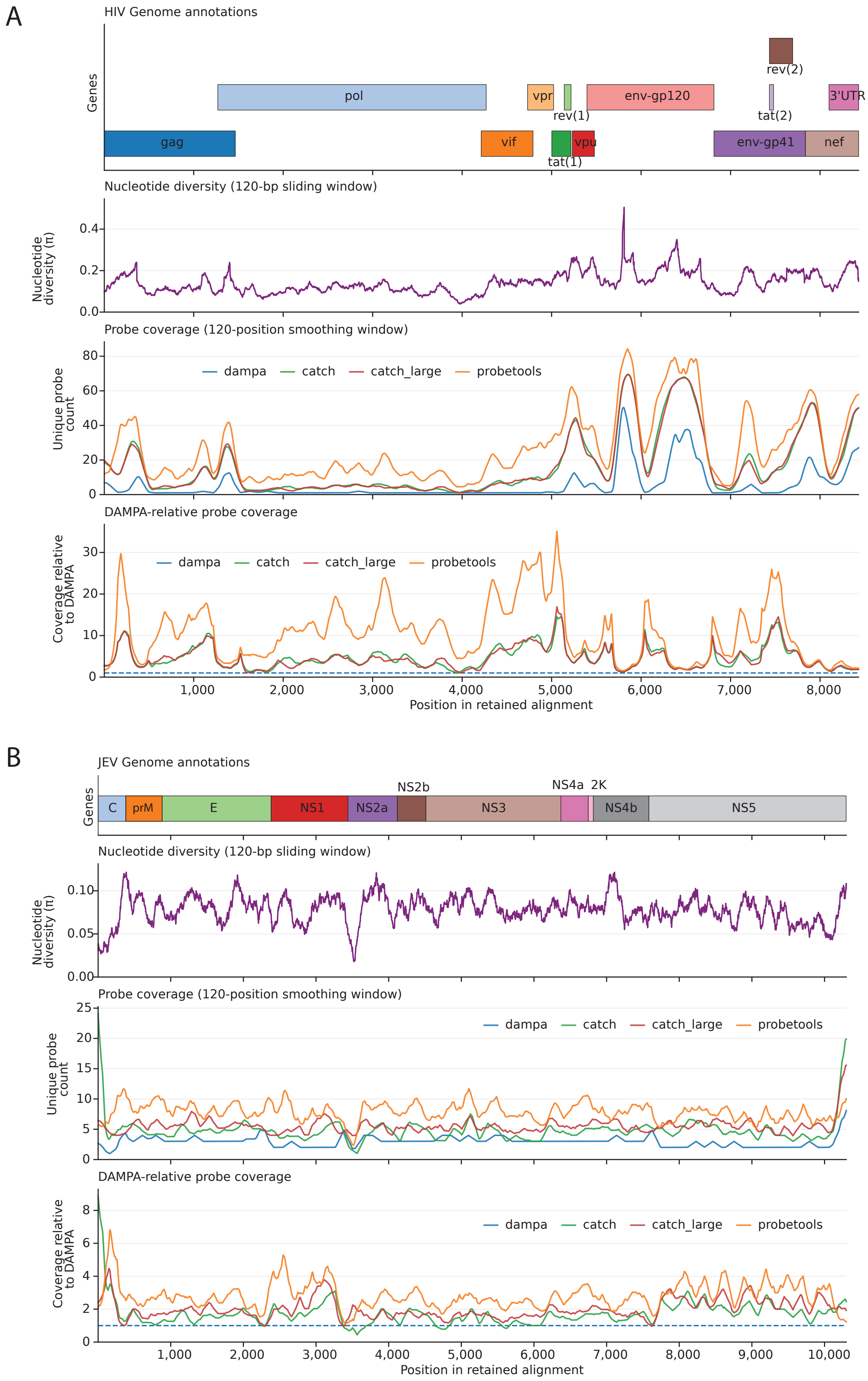


**Supplementary Figure 3** Relationship between probe count from 4 tools and nucleotide diversity using JEV and HIV genome alignments.

A. HIV1 GroupM genomes (72) alignment B. JEV genomes (398) alignment. For each organism, the four panels show the following analyses: i. Gene annotations over the alignment. ii. Nucelotide diversity (π) calculated per alignment position and plotted as a mean over a sliding 120bp window. iii. The number of unique probes predicted to bind to any genome at each position of the alignment. iv. The number of unique probes relative to the number of probes generated by DAMPA, used as a measure of relative composition bias.
