## Supplementary Figure 4 for "DAMPA: compositionally optimised probe panel design for targeted metagenomics"

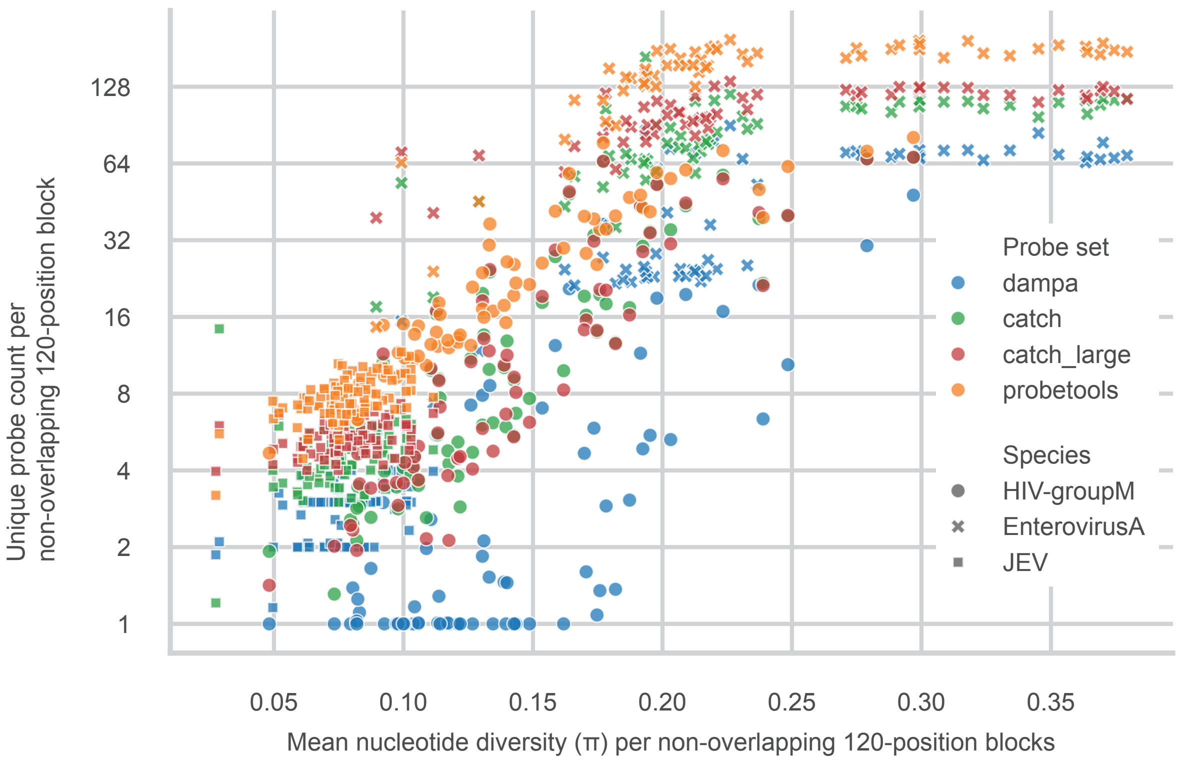


**Supplementary Figure 4.** Relationship between probe count from 4 tools and nucleotide diversity plotted as 120bp datapoints.

Unique probe counts from each of DAMPA, CATCH, CATCH large and Probetools as well as nucleotide diversity (π) were calculated from non-overlapping 120bp alignment sections from Enterovirus A, HIV1 group M and JEV alignments. Each section was plotted for each tool against π. Tools are indicated by colour and species by data point shape.
