## Supplementary Figure 5 for "DAMPA: compositionally optimised probe panel design for targeted metagenomics"

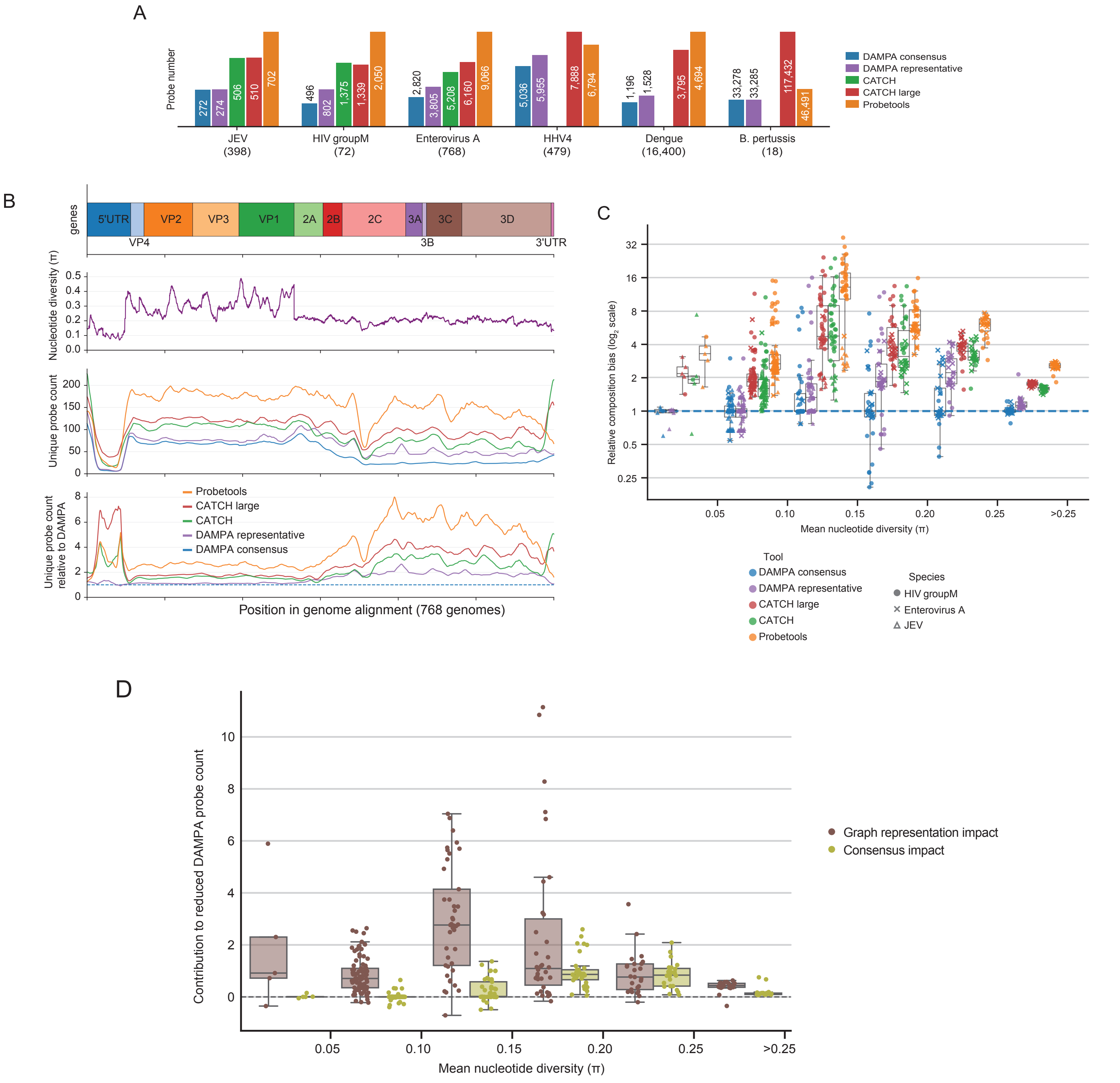


**Supplementary Figure 5.** Performance of DAMPA using representative-based rather than consensus-based probes. A. Probes were designed against the 6 species datasets with a modified version of DAMPA that uses representative sequences from each pangenome graph node rather than the consensus (as described in figure 1B). B. For Enterovirus A, probe counts across the alignment are shown for the four tools and the DAMPA representative-sequence variant. i. Gene annotations over the alignment. ii. Nucelotide diversity (π) calculated per alignment position and plotted as a mean over a sliding 120bp window. iii. The number of unique probes predicted to bind to any genome at each position of the alignment. iv. The number of unique probes relative to the number of probes generated by DAMPA, used as a measure of relative composition bias. C. The relationship between relative composition bias and nucleotide diversity. Mean relative composition bias in each 120bp non-overlapping section of the alignment for each tool was calculated. Relative composition bias counts were binned by nucleotide diversity of the same 120bp section. All counts were normalised by the median of DAMPA consensus probe counts for each bin. D. A comparison of the relative contribution to DAMPA's reduced probe count relative to the closest k-mer based tool. Difference between CATCH and DAMPA-representative relative composition bias represents the improvement attributable to avoiding probe redundancy (brown). The difference between DAMPA representative and consensus relative composition bias represents the improvement attributable to using the consensus sequence to derive probes. The contribution of these two factors is shown across different nucleotide diversity bins.
